# Perioperative Pressure Injury: Bibliometric Analysis

**DOI:** 10.1101/2023.12.04.23299418

**Authors:** Luh Titi Handayani, Moses Glorino Rumambo Pandin, Nursalam

## Abstract

**Background:** The occurrence of pressure injuries in the perioperative period presents a significant clinical hurdle for surgical patients. Surgery is recognized as a risk factor contributing to the onset of pressure injuries, which can manifest shortly after the operation, within a few hours, or up to 72 hours post-surgery. Bibliometric analysis has been carried out to assess publication patterns, rank research studies, and uncover emerging themes in perioperative pressure injury research.

**Method:** Data covering the period from 2015 to 2023 were extracted through the indexing metadata of Scopus and CINAHL using the keywords “pressure injury” OR “pressure ulcer” AND “perioperative.” Bibliometric parameters were extracted, and VOSviewer was employed to obtain bibliometric network and overlay visualization results.

**Result:** The mapping and clustering results from the VOSviewer network and overlay visualization identified various factors, including risk factors, postoperative complications, Electronic Health Record (EHR), prediction models, perioperative leadership, perioperative hypotension, responsibility, skin lesions, positioning, infection, risk assessment, attitude, peripheral nerve injury, and interface injury.

**Conclusion:** The outcomes of mapping, clustering, ranking, and citation analysis of pressure injury in perioperative patients indicate that patient safety is the primary focus. From this main issue, indicators can be developed and transformed into variables that need analysis related to risk assessment, knowledge and attitude, policies, and referrals. The underlying theory in perioperative patient care is the middle-range theory by Myra Estrine Levine, with her esoteric nursing model, Conservation Model of Nursing.

## INTRODUCTION

Pressure Injury, also referred to as pressure ulcer or pressure sore, is characterized by localized damage to the skin and underlying soft tissue. These injuries often manifest over bony prominences or are linked to medical devices and procedures (Lindgren et al., 2005; Mervis & Phillips, 2019). The risk of Pressure Injury is notably associated with surgery, with occurrences possible immediately after the operation, within a few hours, or up to 72 hours in the perioperative phase (Tschannen & Anderson, 2020). The perioperative period encompasses the preoperative, intraoperative, and postoperative stages. Perioperative Pressure Injury is a common challenge in surgical patients, frequently occurring during the intraoperative phase.

Pressure injury is the result of sustained pressure or friction, causing harm to the skin and underlying tissues. Various factors can contribute to the development of pressure injuries, such as the positioning of the patient during surgery, extended surgical duration, physiological shifts in blood circulation and bodily functions throughout the operation, immobility in the preoperative and postoperative stages, utilization of medical devices and supports during the surgical procedure, the patient’s body temperature, ambient room temperature, and their overall health condition. Perioperative pressure injury has a significant impact on patient health and affects postoperative care outcomes, including infection and complications due to microbial entry, delayed wound healing, pain, discomfort, increased costs, prolonged hospital stays, risks of complications such as circulation disorders and deep vein thrombosis, and psychological and quality of life implications.

The incidence of perioperative orthopedic pressure injuries (47, 28.7%) from Braden Scale assessment scores was 19.96 ± 1.81, and the average score of the 3S Intraoperative Risk Assessment Scale was 17.92 ± 3.03. After intraoperative care, 64.0% of patients experience erythema that may resolve, and 6.7% experience stage 1 pressure injuries (Tura et al., 2023). Pressure injury is one of the risk management indicators in healthcare services. Perioperative pressure injury prevention involves preventive actions that the healthcare team can take during the operation period. Nursing actions for pressure injury prevention include risk assessment in the preoperative, intraoperative, and postoperative phases, monitoring positioning, changing positions, maintaining skin moisture, monitoring blood circulation, selecting appropriate intraoperative surfaces, monitoring early signs of pressure injury (risk assessment), and patient and family education. Perioperative pressure injury prevention requires a holistic approach and coordination among healthcare team members involving patients and their families. Preventing perioperative pressure injuries is crucial in caring for patients during and after surgery (Association et al., 2019).

## THEORETICAL FRAMEWORK

The Dever Epidemiological Model proves to be a fitting theoretical foundation for constructing a risk assessment scale aimed at perioperative pressure injuries. This model systematically addresses and categorizes the diverse factors influencing the disease progression, making it apt for investigating the emergence of pressure injuries and their impact on health and disease processes in surgical patients. A literature review on pressure ulcer formation substantiates the importance of employing a theoretical framework to identify risk factors during the perioperative period. Dever’s model acknowledges elements like the surgical setting (specifically, required positioning), the patient’s physiological condition, the use of anesthetic agents, immobility, and the surgical procedure itself as potential contributors to the development of pressure injuries. (Framework Risk Management, n.d.)

A review of several literature sources on perioperative pressure injuries reveals issues regarding the persistently high incidence of pressure injuries during the perioperative phase. The occurrence of pressure injuries during the perioperative phase is considered one of the indicators of healthcare service success, but some incidents are still found in certain hospitals (Bartley and Huntley-Moore, 2022). Factors that may influence the occurrence of pressure injuries are related to concepts such as biopsychosocial, Health Belief, prevention, Transtheoretical Model, Risk Assessment Model, and Evidence-Based Nursing (EBN).

The biopsychosocial model considers the interaction of biological, psychological, and social factors in health and disease. This model indicates that pressure ulcers can be influenced by physical factors (such as tissue ischemia and friction forces) and psychosocial factors (such as patient mobility, pain perception, and mental health). Interventions must address the multifaceted nature of pressure ulcer development. An individual’s Health Belief Model about health risks and the perceived benefits of taking action to reduce those risks would involve understanding patients’ beliefs about being susceptible to developing pressure ulcers and their perceptions of the effectiveness of preventive measures.

Preventing pressure injuries may involve understanding key elements contributing to the development of pressure sores. This can include factors such as pressure intensity and duration, tissue tolerance, and the effectiveness of preventive interventions. A behavior change process can be implemented to pressure injury prevention. Preventing pressure injuries in the perioperative environment involves a dynamic and adaptive process. Understanding the complexity of the healthcare system and the interactions among patients, healthcare providers, and interventions is crucial for effective prevention. Risk Assessment Models are used to predict the likelihood of pressure injury development. These models often include factors such as age, mobility, nutritional status, and comorbidities. Understanding and applying these models can assist healthcare providers in identifying high-risk patients and implementing targeted prevention strategies (Javan Biparva et al., 2023).

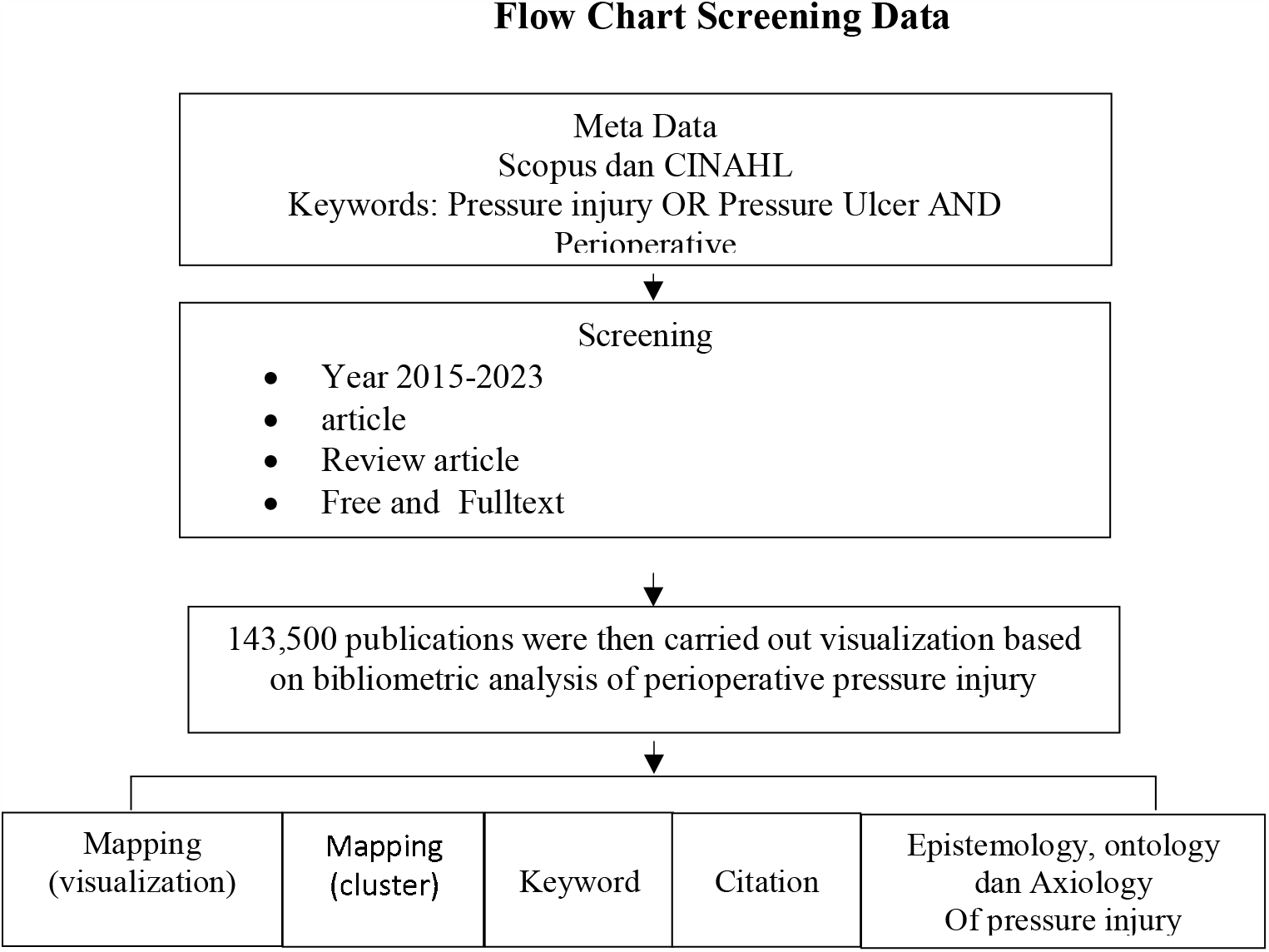

Risk Assessment Model to predict the likelihood of developing pressure ulcers. These models often include factors such as age, mobility, nutritional status, and comorbidities. Understanding and applying these models can help healthcare providers identify high-risk patients and implement targeted prevention strategies. Pressure ulcer prevention in the perioperative setting can also be based on evidence-based practice. It involves integrating the best available evidence, clinical expertise, and patient preferences to make informed decisions about patient care. It is important to note that the choice of theoretical framework depends on the context, goals, and specific characteristics of the healthcare setting. Integrating multiple theoretical frameworks may provide a more comprehensive understanding of perioperative pressure ulcer prevention and management.

Research Questions

1. How is pressure injury research mapped (visualized)?
2. How does perioperative pressure injury cluster?
3. From which countries are researchers on perioperative pressure injuries?
4. What is the perioperative pressure injury article with the highest number of citations?

What is the review of the philosophy of pressure injury from ontology, epistemology, and axiology?

## METHODOLOGY

Bibliometric analysis is a method based on understanding that involves quantitative assessment of the literature in a particular field. Phases in bibliometric analysis include keyword identification, literature search, data extraction, data analysis, data visualization, and interpretation of results. Keyword identification was carried out using the keywords “pressure injury” OR “pressure ulcer” AND “perioperative” database from 2015 - 2023 from the Scopus and CINAHL search engines. Search and selection of studies refers to reviews and articles in Indonesian and English. Data extraction was carried out and 143,500 metadata were obtained. The data that has been obtained is then processed with VOSviewer software version 1.6.20. VOSviewer data analysis is used to carry out co-occurrence analysis, a valuable technique for identifying patterns and relationships in document networks.

Bibliometric analysis is the quantification of academic (written) output and its perception. Classification of community and institutional performance based on bibliometric indicators is presented in the form of rankings. Statistics produce indirect clues about the quality of academic performance of communities, institutions, and countries (Ball, 2017). Bibliometrics is the quantitative analysis of scientific publication and citation data, aimed at providing insight into the value and impact of published research. Bibliometry can be used to 1) show the impact of research, 2) Identify collaborators and experts, 3) Identify emerging research trends, 4) Provide information about research priorities, and 5) Select the most relevant journals for publication.

## RESULTS AND DISCUSSION

### Mapping (Visualization) Penelitian Pressure Injury

Network Visualization VOSViewer Perioperative Pressure Injury Based on Figure 1, several variables were found that were associated with the incidence of perioperative pressure injury. Based on the VOSViewer visualization overlay, several variables and indicators of perioperative pressure injury that can be developed (mapping for the 2015-2023 period) include, identification of risk factors, postoperative complications, Electronic Health Record (HER), prediction model, perioperative leader, perioperative hypotension, responsibility, skin lesions, positioning, infection, risk assessment, attitude, peripheral nerve injury, interface injury.

**Figure 1.**
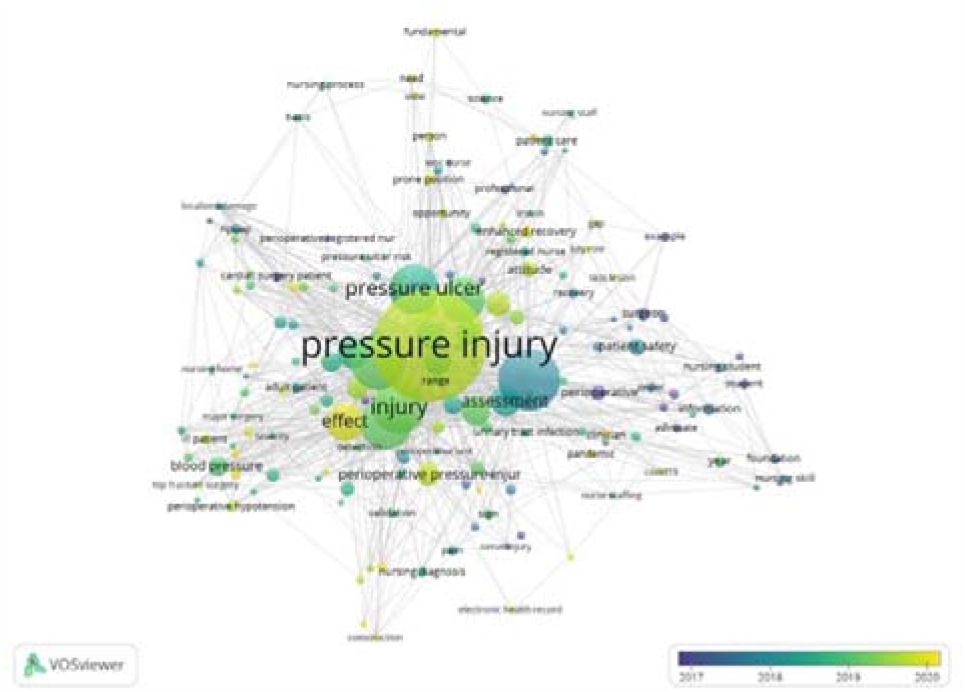
Network Visualization VOSViewer Pressure Injury Perioperative

The results in pressure injury mapping will present the results of the VOSviewer bibliometric analysis and metadata, namely 1) articles related to pressure injury based on the author, 2) selecting relevant journals, 3) searching for information about research priorities, 4) providing information Research trends of perioperative pressure injury. The results of the research trend analysis used the co-occurrence feature in VOSviewer to find keywords in the research title and abstract. Keywords are grouped according to the year the article was published.

### Perioperative Pressure Injury Cluster

Table 1 describes the clusters of perioperative pressure injury, cluster one (1) contains guidelines and assessment, cluster two (2) concerns physical influences and pathophysiology, cluster three (3) concerns referrals and regulations, cluster four (4) identifies risk factors, cluster five (5) nursing treatment, cluster six (6) individuals at risk of the impact of surgery, cluster seven (7) pressure injury, cluster eight (8) regarding the knowledge and attitudes of nurses (surgical team), cluster nine (9) regarding the impact and causes of pressure injury, cluster ten (10) regarding environmental support and facilities and infrastructure, cluster eleven (11) regarding hypothermia.

**Table 1.**
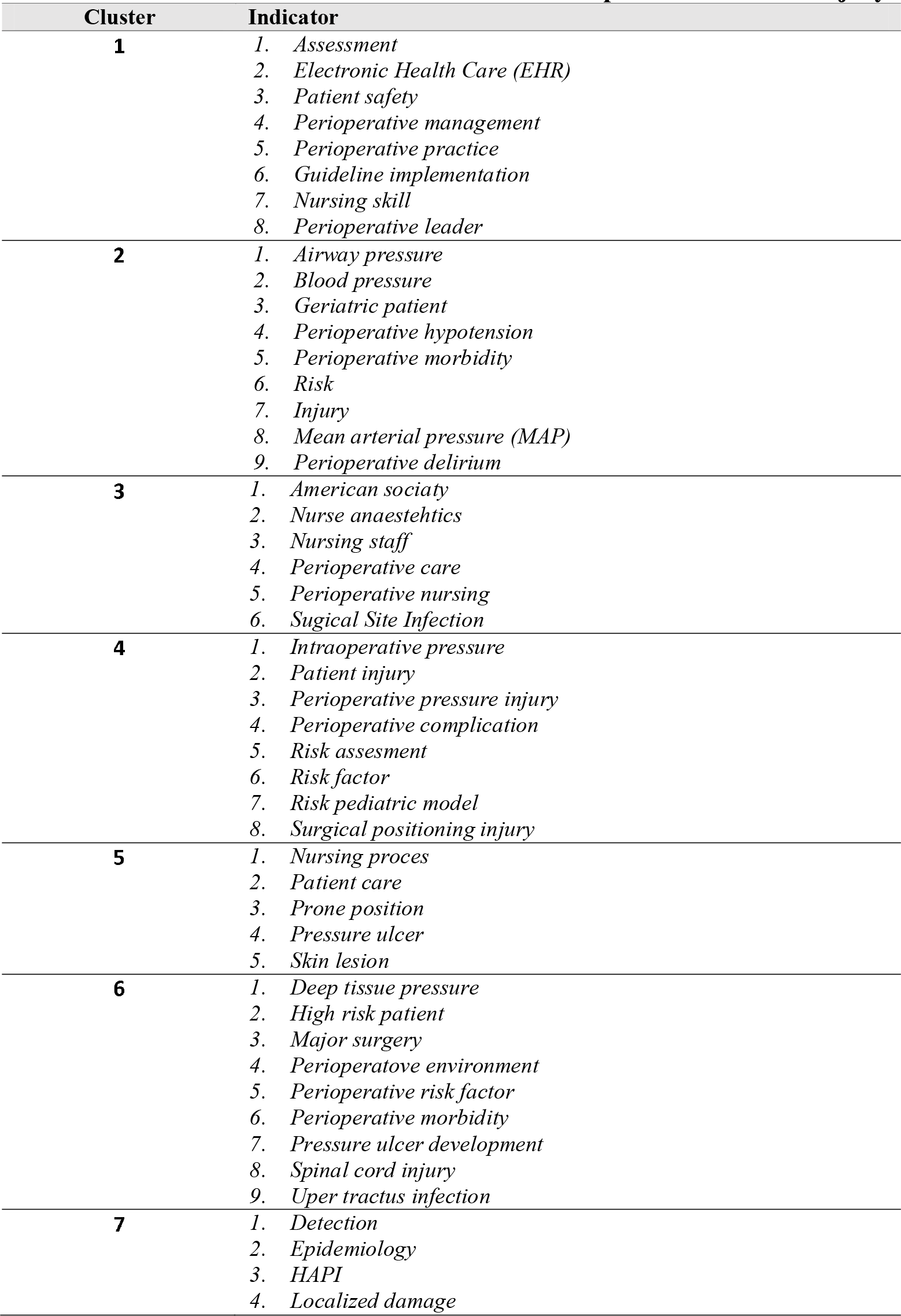
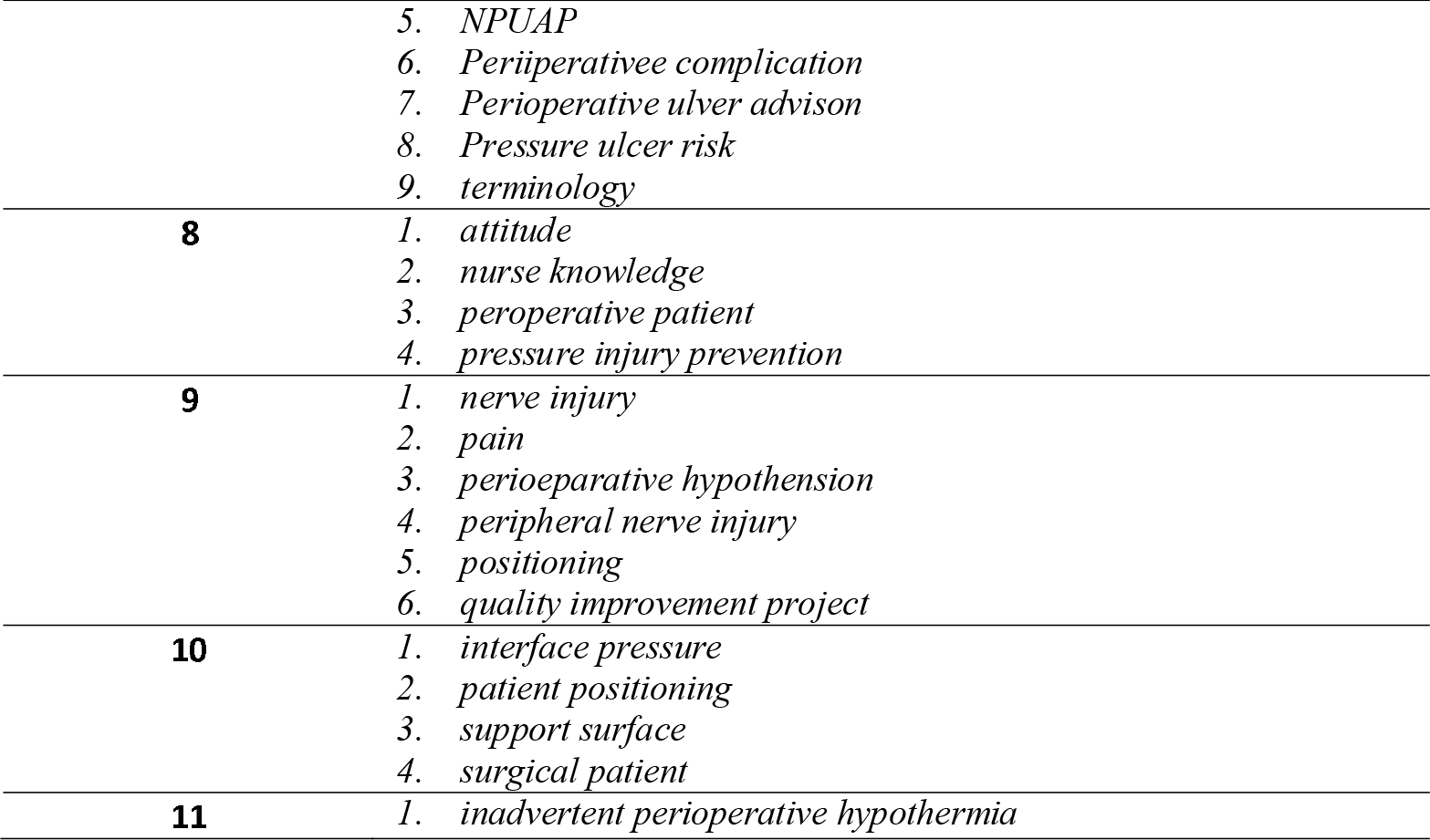
VOSviewer Clusters on Perioperative Pressure Injury.

Table 2 illustrates the top ten (10) publications on the theme of perioperative pressure injury based on citations and searches by keywords. The terms written by the author from the 10 sequences of citations on the topic of perioperative pressure injury above explain the topics of acquired pressure injury, patient safety, risk assessment tools, responsibility, knowledge, and attitude of the operating room team. Journal (1) describes Healthcare strategies and efforts to improve patient outcomes, journal (2) risk factors that may contribute to the development of pressure ulcers, journal (3) risk factors for pressure injuries in the perioperative patient, Journal (4) understanding emerged that the operating theater nurse, journal (5) indicates the continued need for further resource allocation into PU prevention and management, journal (6) different aspects of caring in perioperative practice, Journal (7) Constantly managing risk and preventing the OT patient from harms, Journal (8) predicting surgical risk factors, risk assessment (9) Knowledge Practice Nurse Pressure ulcer prevention improving the safety of patients from pressure injury perioperative, Journal (10) The scale of risk assessment wound and injuries perioperative nursing.

**Table 2.** Ten articles about Perioperative Pressure Injury based on keywords and conclusions.

| No | Title | Article Type | Keyword | Conclusi |
| --- | --- | --- | --- | --- |
| 1 | The relationships of nurse staffing level and work environment with patient adverse events | original article | Korea, nurse staffing, patient adverse events, patient outcomes, work environment | Enhancing patient outcomes requires the implementation of healthcare strategies and initiatives aimed at adjusting appropriate nurse staffing levels and creating improved work environments for nursing staff. (Cho et al., 2016) |
| 2 | Pressure ulcers: factors contributing to their development in the OR | review article | pressure ulcer, risk factors, HAPU, never event. | The study uncovered recurring risk factors that might play a role in the formation of pressure ulcers in patients. (Engels et al., 2016) |
| 3 | Back to basics: preventing perioperative pressure injuries | review article | pressure ulcer, pressure injury, HAPI, patient positioning, preoperative assessment. | The factors that pose a risk for pressure injuries in patients undergoing perioperative care. |
|  |  |  |  | (Spruce, 2017) |
| 4 | Responsibility for patient care in perioperative practice | review article | ethic, hermeneutic text interpretation, operating theatre nurse, perioperative nursing, responsibility | The ethical value of confirming the patient involves validating their identity or ensuring their identity is accurate (Moore et al., 2019) |
| 5 | The prevalence of pressure ulcers in Europe, what does the European data tell us: a systematic review | original article | cross-sectional studies, epidemiology, Europe, prevalence, pressure injury, pressure sore, pressure ulcers | These imply that more funding should be set out for pressure ulcer management and prevention.. (Moore et al., 2019) |
| 6 | Making the invisible visible–operating theatre nurses' perceptions of caring in perioperative practice | original article | care; operating theatre nurse; perioperative nursing. | Various elements of providing care in perioperative practice were explored or discussed. (Blomberg et al., 2015) |
| 7 | Enhancing patient safety in the operating theatre: from the perspective of experienced operating theatre nurses | review article | patient safety, operating theatre, operating theatre nurses, perioperative nursing, phenomenology, interviews | Continuously mitigating risks and safeguarding the patient in the operating room to ensure patient safety (Ingvarsdottir and Halldorsdottir, 2018) |
| 8 | A prediction tool for hospital-acquired pressure ulcers among surgical patients: Surgical pressure ulcer risk score | original article | nosocomial, pressure injury, surgery, surgical risk factors, risk assessment | Identifying the risk of pressure ulcers before surgery. (Aloweni et al., 2019) |
| 9 | Factors related to knowledge, attitude, and practice of nurses in intensive care unit in the area of pressure ulcer prevention: A multicenter study | original article | Pressure ulcers Bed sores Attitude Knowledge Practice Nurse Pressure ulcer prevention | The degree of attitude and knowledge that nurses possess, along with the comparatively good practices that they display when it comes to pressure ulcer prevention, highlight how important it is to improve patient safety.(Khojastehfar et al., 2020) |
| 10 | Assessment scale of risk for surgical |  | Perioperative Nursing; Intraoperative Period; Nursing Care; Wounds | Even though the scale is a legitimate and trustworthy tool, more |
|  | positioning injuries |  | and Injuries; Risk Assessment; Patient Positioning. | investigation is needed to determine whether or not it can be used in clinical situations.(Lopes et al., 2016) |

### Perioperative Pressure Injury Articles With the Highest Number of Citations

Table 3 depicts twenty-five (25) major publications on the theme of perioperative pressure injury based on citations. The terms written by the author from the 25 series of citations on the topic of perioperative pressure injury above explain the topics of acquired pressure injury, patient safety, risk assessment tools, responsibility, knowledge, and attitude of the operating room team.

**Table 3.**
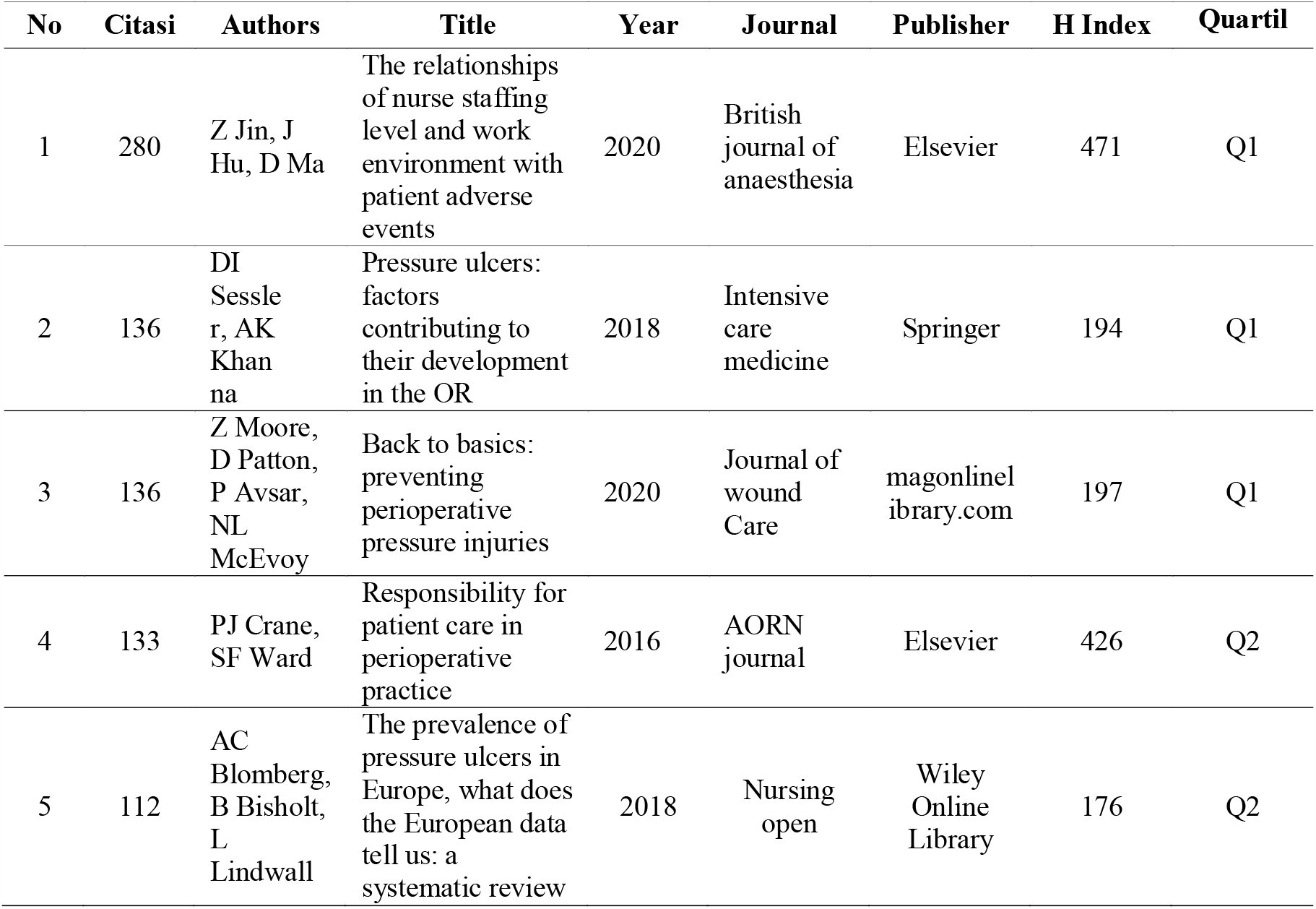
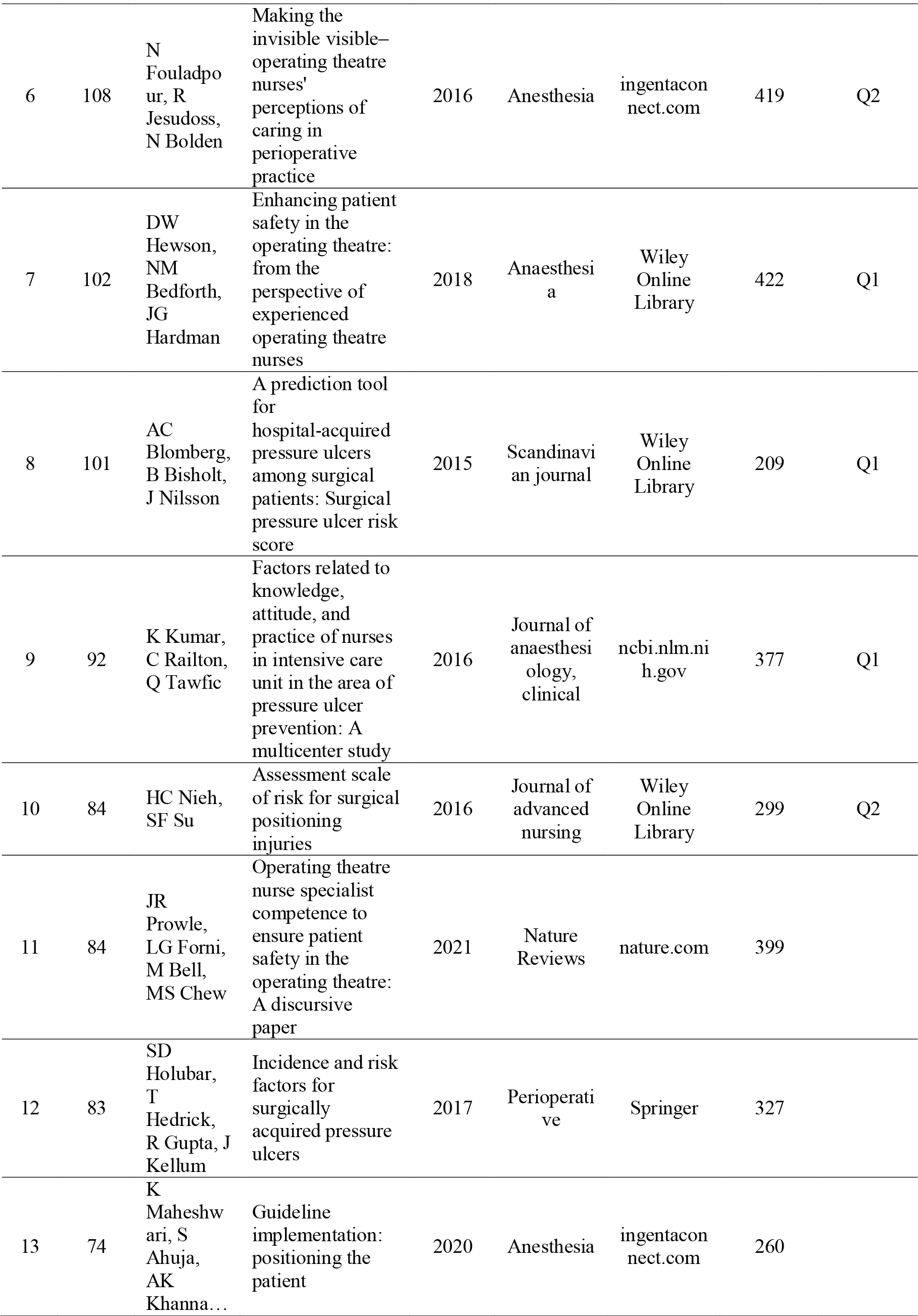
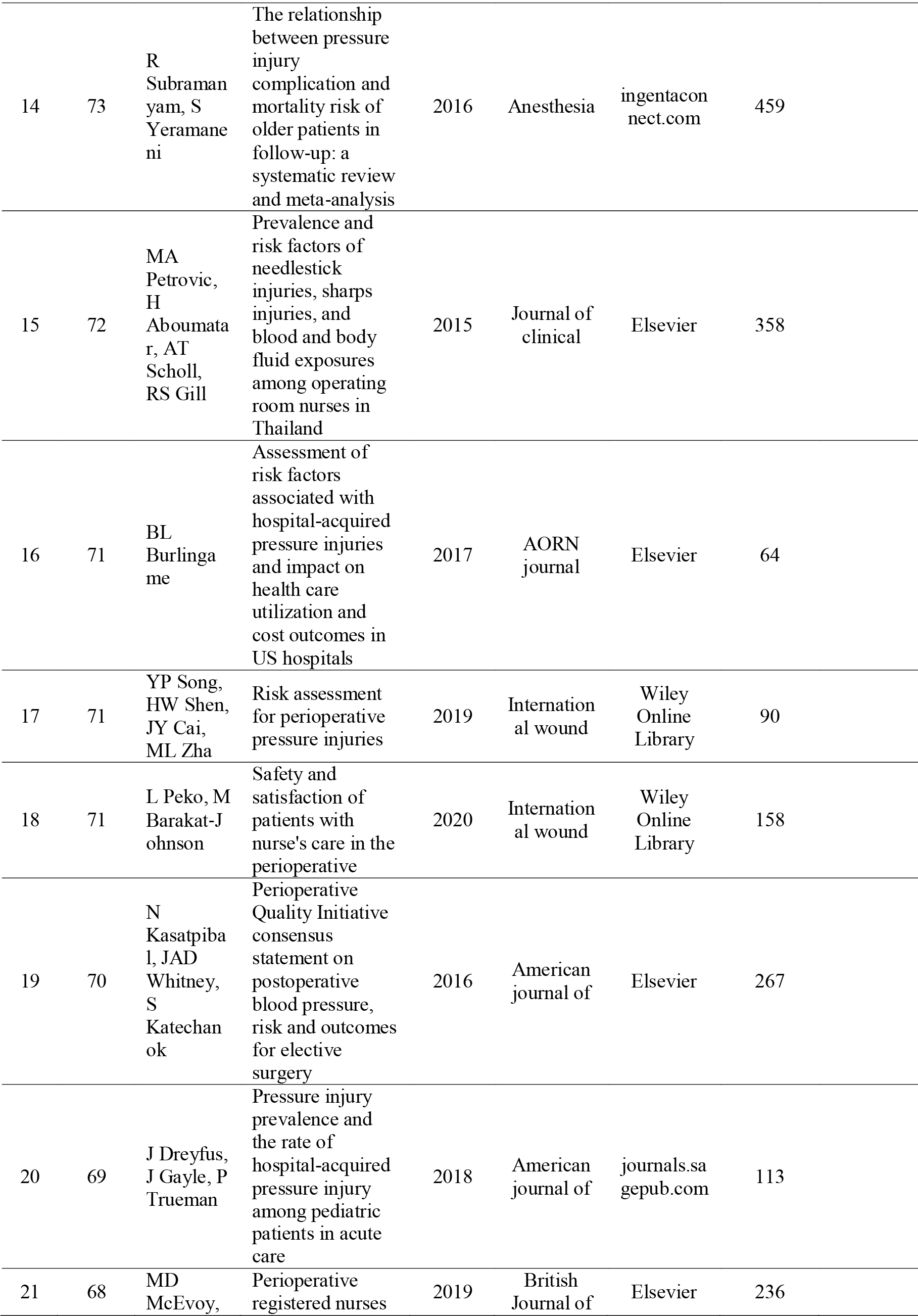
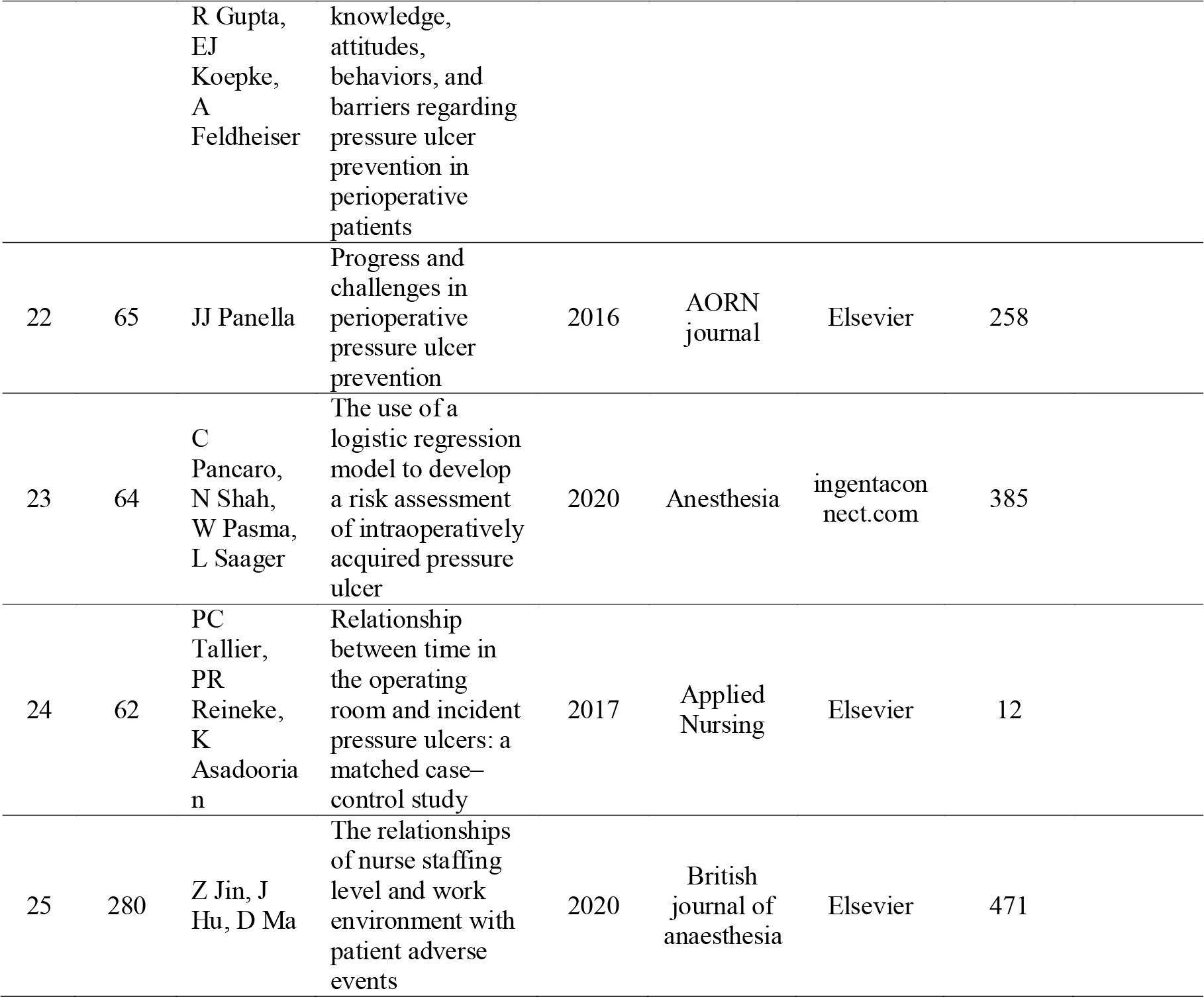
Twenty-five (25) Articles Related to Pressure Injury Research in Nursing Disciplines Author.

### Review of the ontology, epistemology, and axiology of the concept of pressure injury

The Four Principles of Conservation Model, often known as the esoteric nursing model “Nursing Conservation Model” developed by nursing theorist Myra Estrine Levine, is well-known. By preserving energy and resources, this model promotes patients’ optimum health and well-being and provides a foundation for nursing practice. The conservation model of nursing developed by Myra Levine emphasizes the nurse’s responsibility to assist patients in coping with health issues and protecting their social, emotional, and physical resources. This paradigm promotes a comprehensive approach to nursing care that takes the patient’s psychological and social needs into account in addition to their physical health. (Mc Ewn and M. Wils, 2019).

### Ontology

The First of Four Conservation Principles: Aiming to direct the provision of nursing interventions that can affect a person’s response and enhance their state of well-being, the Model is a model for nursing care. 2) Individual energy conservation means striking a balance between energy intake and output to prevent overexertion while getting enough sleep. adequate activity, diet, and 3) Preservation of individual structural integrity is the process of preserving or repairing the bodily structure, averting harm, and quickening the healing process. Examples of this include assisting patients with range-of-motion (ROM) exercises and maintaining good personal hygiene. 4) Maintaining each person’s integrity as an individual and acknowledging them as people who aspire to respect; acknowledgment, self-awareness, and autonomy 5) When the patient is identified as a member of a family, community, religious organization, etc., social integrity is preserved (Wijaya et al., 2022).

Applying ontological philosophy to pressure injuries in the perioperative setting involves considering the ontological viewpoint on the essence of existence and reality during surgical procedures. In this scenario, the ontological approach requires a deep comprehension of pressure injuries as tangible phenomena, their connection to the patient’s reality, and the interpretation of these experiences within an ontological framework. This philosophical approach may also extend to broader considerations regarding the existence and relationships among patients, nurses, and the perioperative environment, aiming for a comprehensive understanding of the ontological implications of pressure injuries in this context.

### Epistemology

Epistemology in Myra Estrin Levine’s theory in nursing reflects a holistic approach to understanding nursing knowledge. It integrates practice experience, science, and a deep understanding of the patient to provide care that is effective and focuses on the patient’s individual needs. The epistemological approach to perioperative pressure injuries involves how we understand and acquire knowledge about this phenomenon during the surgical period. In this context, epistemology encompasses questions about how we know, how knowledge about pressure injuries is obtained, and how this knowledge can be considered valid and reliable.

In terms of the epistemological perspective on perioperative pressure injuries, it is essential to consider various sources of knowledge, including scientific evidence, clinical experience, and patient understanding. Additionally, epistemology involves considerations of the research methodologies used to gather information about pressure injuries and the extent to which such knowledge can be relied upon. This epistemological viewpoint reflects a philosophical approach to the understanding and construction of knowledge about perioperative pressure injuries, considering how knowledge is acquired, tested, and interpreted within the context of healthcare practice.

### Axiology

The context of Myra Estrin Levine’s theory in nursing emphasizes the importance of values and ethics in her nursing practice by applying: 1) the value of patient safety as the main value in patient safety and welfare, 2) professional ethics includes professional ethics that regulate nurses’ behavior from justice, patient autonomy, and avoiding conflicts of interest, 3) Empathy and caring values emphasize the importance of empathy and caring in the nurse-patient relationship, 4) Sustainability Principles include the principles of sustainability in care, 5) Humanistic values reflect a humanistic approach to health care. This includes respect for human uniqueness and dignity as well as recognition of patient values, beliefs, and preferences.

Although the term axiology is rarely used in a medical or healthcare context, we can try to understand an ethical or values perspective on perioperative pressure injury. In this context, axiology can refer to the consideration of the values and ethics underlying the management of perioperative pressure injuries. Some ethical considerations that may arise include: 1) Prevention: Ethical values encourage maximum effort in preventing perioperative pressure injuries as a form of patient protection, 2) Patient Empowerment: Healthcare ethics often emphasizes the importance of empowering patients with appropriate information and understanding. adequate regarding the risk of pressure injuries as well as ways to prevent them, 3) Justice: Ethical considerations regarding the distribution of health care resources and attention may arise, especially when it comes to preventing pressure injuries in vulnerable populations, 4) Quality of Care: Ethical values that focus on providing quality and safe health care may guide efforts to reduce perioperative pressure injuries, 5) Openness and Transparency: Organizational ethics may encourage openness and transparency regarding pressure injury incidents, as well as efforts to continually improve standards of care, 6) An axiological view in the context of pressure injuries Perioperative care may involve moral considerations and ethical values underlying health care decisions and actions to protect and promote patient well-being.

## CONCLUSIONS

The results of mapping and clusters as well as the ranking and citation of the term pressure injury in perioperative patients illustrate that the main topic is patient safety. From the main problem, the indicators obtained can be developed into variables that need to be analyzed related to risk assessment, knowledge and attitudes, policies, and references. The underlying theory in perioperative patient care is the middle ring theory of Myra Estrine Levine with her esoteric nursing model “Nursing Conservation Model”.

## Data Availability

All data produced are available online at

